# Single-Center Hospital and Outpatient Opioid Use for Lower Extremity Arterial Disease

**DOI:** 10.1101/2024.04.22.24306170

**Authors:** Xuanjia Fan, Nicholas M. Graziane, Maria C. Ramirez, Salvatore L. Stella, Prasanna Karunanayaka, Victor Ruiz-Velasco, Sanjib Adhikary, Flohr Flohr

## Abstract

**Introduction:** The pain associated with lower extremity arterial disease is difficult to treat, even with lower extremity revascularization. We sought to evaluate in-hospital and post-operative opioid usage in patients with different disease severities and treatments for lower extremity vascular disease.

**Methods:** A retrospective review was performed for all hospital encounters for patients with an International Classification of Diseases (ICD) code consistent with lower extremity arterial disease admitted to single center between January 2018 and March 2023. Encounters were subdivided according to patient’s disease severity, treatment type as designated by Current Procedural Terminology (CPT) code and comorbid diagnosis of diabetes mellitus. These groups were analyzed for in-hospital opioid use frequency and dosage. Encounters for patients admitted with a secondary diagnosis of lower extremity atherosclerotic disease were included as the control group (CON). A total of 438 patients represented by all the analyzed encounters were then reviewed for number and type of vascular procedures performed as well as opioid use in the outpatient setting for one year.

**Results:** Critical limb ischemia (CLI) encounters were more likely to use opioids as compared CON and peripheral arterial disease (PAD) without rest pain, ulcer or gangrene groups (CLI 67.9% versus CON 52.1%, *p* < 0.001 and CLI 67.9% versus PAD 50.2%, *p* < 0.001). Opioid use was also more common in encounters for gangrene and groups treated with revascularization (REVASC) and amputation (AMP) as compared to CON (gangrene 74.5% versus CON 52.1%, *p* < 0.01; REVASC 58.3% versus CON 52.1%, *p* =0.01; and AMP 72.3% versus CON 52.1%, *p* < 0.01). Significantly increased opioid doses per day (MME/day) were not noted for any of the investigated groups as compared to CON. In the outpatient setting, 186 (42.5%) patients were using opioids one month after the most recent vascular intervention. By one year, 31 (7.1%) patients were still using opioids. No differences in opioid usage were noted for patients undergoing single versus multiple vascular interventions at one year. Patients undergoing certain vascular surgery procedures were more likely to be using opioids at one year.

**Conclusion:** Patients with CLI and gangrene as well as those undergoing vascular treatment have a greater frequency of opioid use during hospital encounters as compared those patients with less severe disease and undergoing conservative management, respectively. However, these findings do not equate to higher doses of opioid used during hospitalization. Patients undergoing multiple vascular procedures are not more likely to be using opioid long-term (at one year) as compared to those patients treated with single vascular procedures.

**ARTICLE HIGHLIGHTS:** *Type of Research:* A single-center, retrospective review

*Key Findings:* Patients with critical limb ischemia during hospitalization use opioids more frequently than patients with less severe vascular disease. These findings do not equate to higher doses of opioid used during hospitalization. Patients with lower extremity arterial disease undergoing multiple vascular procedures are not more likely to be using opioids long-term (at one year) as compared to patients treated with single vascular procedures.

*Take Home Message:* The pain associated with lower extremity arterial occlusive disease often requires treatment with opioids, however, disease severity and numerous treatments do not equate with the need for increased or prolonged dosage.

## INTRODUCTION

Short of revascularization, the pain associated with limb ischemia is difficult to treat [1,2]. Even with revascularization, patients are often left with augmented post-surgical pain as a result of long incisions made in unhealthy tissue and swelling associated with reperfusion [3]. Patients who are not candidates for revascularization and require amputation for limb ischemia also have unresolved pain exacerbated by phantom limb syndrome [4,5]. Patients dealing with limb ischemia are often discouraged from using nonsteroidal anti-inflammatory drugs because of the potential for worsening chronic renal insufficiency and potentiating bleeding risk [6]. Regional anesthetic options for the treatment of pain are temporary and limited [7]. These treatment constraints leave opioids as the most frequently used analgesics for ischemic limbs. Opioids, while initially efficacious, eventually lose their effectiveness requiring increasing dosages, resulting in patient tolerance and dependence [8,9].

Surgeon self-analysis over the past decade has revealed that patients undergoing vascular surgery are at high risk of developing opioid dependence with approximately 10% of patients using opioids beyond three months [10]. Recently, vascular surgeons sought to attribute complications from vascular surgery with prolonged opioid use and they found over 25% of patients undergoing lower extremity revascularization were still using opioids beyond six months of their original intervention with the strongest predictor of long-term use being prior opioid use [11]. Little, if any, research looks at in-hospital opioid use in patients with lower extremity vascular disease and follows those patients for continued opioid use. Given the need to minimize opioids for treating chronic pain, performing this research will help identify current in-hospital treatment strategies for patients with limb ischemia and whether alternative treatment options should be considered to reduce opioid use.

This study sought to determine 1) how frequently admission for lower extremity arterial disease is treated with opioids, 2) how vascular disease severity affected in-hospital opioid use, and 3) how treatment with either revascularization or amputation as opposed to patients admitted with a secondary diagnosis of lower extremity atherosclerotic disease (controls) effect the rate of in-hospital opioid use. We hypothesized that opioid use is increased with disease severity, with a secondary diagnosis of diabetes and with vascular disease treatment as opposed to controls. Finally, using institutional data, this study sought to determine if multiple hospital admissions for multiple vascular procedures (both revascularization and amputation) led to increased post-operative opioid use. We hypothesized that patients undergoing more vascular procedures were more likely to continue using opioids in the post-operative period.

## MATERIALS AND METHODS

### Institutional data, in-hospital opioid use

All hospital admissions for patients with a diagnosis of lower extremity vascular disease were reviewed for Penn State Milton S. Hershey Medical Center between January 2018 and March 2023. Permission for this retrospective review was granted by the Penn State College of Medicine Institutional Review Board, IRB protocol 22745. Hospital encounters for review were identified by International Classification of Diseases (ICD) codes designating patients with lower extremity atherosclerotic disease or lower extremity diabetic peripheral angiopathy which are listed in **Supplemental Table 1**. Encounters for patients with ICD codes designating atherosclerotic disease in an “other” extremity or an “unspecified” extremity were excluded from this analysis.

Encounters were further subcategorized by patient vascular disease using listed ICD diagnosis codes. Encounters for patients having peripheral arterial occlusive disease (PAD) were designated if the diagnosis code did not include rest pain, ulceration or gangrene or if the code listed claudication. Encounters for patients were subcategorized as critical limb ischemia (CLI) if the diagnosis code listed included rest pain, ulceration or gangrene. Encounters with CLI and PAD were further subcategorized into those with an underlying diagnosis of diabetes mellitus (DM). Separating patients with PAD and critical limb ischemia CLI based on DM diagnosis allows for the examination of opioid use patterns in more homogeneous subgroups, considering potential differences in pain profiles, treatment strategies, and clinical outcomes.

Encounters were also subcategorized by intervention during hospital admission. Patients admitted with a secondary diagnosis of lower extremity atherosclerotic disease were control subjects (CON). These encounters included patients with a vascular diagnosis admitted for reasons other than vascular disease. The encounter was categorized as being for revascularization (REVASC) if the admitted patient had a current procedure terminology (CPT) code designating a revascularization procedure. These procedures included angiogram +/- angioplasty, stenting, embolectomy, aortobifemoral or aortobi-iliac bypass, endarterectomy, and lower extremity bypass. The encounter was categorized as being for major amputation (AMP) if an associated CPT code designated below or above knee amputation. Encounters for patients undergoing isolated toe or transmetatarsal amputations during hospitalization were grouped with REVASC group. Vascular surgery CPT codes identified and associated with CON, REVASC and AMP groups are listed in **Supplemental Table 2**.

CON was compared to patients admitted for PAD and to patients admitted for CLI. Groups were compared based on opioid usage and amount used during hospitalization. Encounters by vascular disease ICD code designation (as having claudication, rest pain, gangrene, or isolated atherosclerotic disease without the aforementioned complaints) were evaluated for opioid use and amount used during hospitalization. CON was compared to encounters representing patients undergoing REVASC and encounters representing patients undergoing AMP for opioid usage and amount during hospitalization.

Demographics assessed for the aforementioned groups include average age, male gender, average days admitted during the encounter. Hospitalization encounters were considered positive for opioids if the following medications were listed as being given during the admission: hydromorphone, Dilaudid, morphine, MS Contin, methadone, Dolophine, meperidine, Demerol, fentanyl, Duragesic, Sublimaze, oxycodone, Percocet, hydrocodone, Vicodin, and codeine. Encounters with opioids given only at the time of surgical intervention were not considered positive for opioids. Average daily morphine milligram equivalents were calculated for the encounters.

### Institutional data, outpatient opioid use

Electronic medical records for individuals with PAD and CLI having multiple hospital admissions during the selected time period were reviewed. Patient demographics including age (as of January 2018), gender, and most recent survival status were collected. The vascular surgical interventions were categorized as angiogram +/- angioplasty, stenting, embolectomy, aorto-bi-femoral or aorto-bi-iliac bypass, endarterectomy, lower extremity bypass, toe amputation, transmetatarsal amputation, below knee amputation or above knee amputation. Patients were categorized as REVASC or AMP.

In order to better evaluate whether the number vascular interventions were contributing to long-term opioid usage, patients with multiple vascular interventions during the period of inquiry were evaluated. The date of the patient’s last vascular surgical intervention was noted, and subsequent outpatient surgical evaluations were reviewed for patient narcotic usage. Patients having narcotics listed on their outpatient surgical records beyond one month after their last vascular surgical intervention were considered chronic narcotic users. If the patient did not have narcotics listed on their outpatient surgical records beyond one month, they were considered not chronic narcotic users. For those patients with narcotics listed greater than one month beyond their last vascular surgery date, the number of months positive for narcotics use was recorded. For example, patients with outpatient surgical records for one month and three months post vascular surgery were considered to be using narcotics chronically for three months, even though no outpatient surgical records for two months were available to review. If the same patient was seen five months post vascular surgery and no narcotics were listed on the fifth month outpatient records, the patient was considered a chronic narcotic user for three months only.

### Statistical analysis

All variables were summarized with frequencies and percentages or medians with interquartile ranges (IQR). Variables were compared using Wilcoxon and chi-squared tests for continuous and categorical variables, respectively. Pearson’s correlation coefficient was used to determine correlation between continuous variables and reported with *r* values. Statistical significance was set at a *p*-value of < 0.05. Analyses were performed using R version 4.3.2 statistical package, 2023 (Vienna, Austria) or GraphPad Prism software version 10.0.2.

## RESULTS

### In-hospital opioid use for vascular encounters

#### A total of 1359 in-patient hospital encounters

(1043 patients) were identified during the five-year period (Jan. 2018-March 2023). Demographics for CON, vascular disease severity (PAD and CLI), vascular signs/symptoms on presentation (claudication, rest pain, ulcer, and gangrene) and vascular treatment groups (REVASC and AMP) are listed (**Table 1**). Ages for each group were similar to CON with exception of REVASC encounters (CON: 69 (62-77) years; REVASC: 67 (60-74); *p* < 0.01, Wilcoxon test) (**Table 1**). Sex distribution was similar for all groups of encounters as compared to CON (PAD vs. CON: χ^2^ = 0.0005, df =1; CLI vs CON: χ^2^ = 0.07, df =1; chi-squared test) (**Table 1**). PAD, claudication, rest pain, and REVASC encounters had shorter admissions as compared to CON (PAD vs. CON: *W* = 125082, *p* < 0.001; Claudication vs. CON: *W* = 38593, *p* < 0.01; Rest pain vs CON: *W* = 45795, *p* < 0.01; REVASC vs. CON: *W* = 164888, *p* = 0.01; Wilcoxon test) (**Table 1**). Gangrene and AMP encounters had longer admissions as compared to CON (Gangrene vs. CON: *W* = 56814, *p* = 0.03; AMP vs. CON: *W* = 69420, *p* < 0.01; Wilcoxon test) (**Table 1**).

**Table 1.** Institutional data separated by vascular disease severity, vascular signs/symptoms on presentation and vascular treatment. Percentage of hospital admissions positive for opioids noted across “+ opioid” row. *p* values as compared to CON. \**p* < 0.05; **p < 0.001. Abbreviations: LOS = length of stay.

| Group<br>n (%) | Vascular Disease Severity |  |  | Vascular Signs/Symptoms on Presentation |  |  |  | Vascular Treatment |  |
| --- | --- | --- | --- | --- | --- | --- | --- | --- | --- |
|  | CON<br>793 (58.4%) | PAD<br>242 (17.8%) | CLI<br>324 (23.8%) | Claudication<br>64 (5.5%) | Rest pain<br>97 (8.4%) | Ulcer<br>41 (3.54%) | Gangrene<br>161 (13.9%) | REVASC<br>379 (26.8%) | AMP<br>242 (17.1%) |
| Age in years<br>Median (IQR)<br>*p* value | 69 (62-77) | 69.5 (62-75)<br>0.37 | 68 (61-77)<br>0.5 | 68 (63-72)<br>0.38 | 66 (61-74)<br>0.11 | 69 (62-80)<br>0.32 | 67 (61-77)<br>0.45 | 67 (60-74)*<br>< **0.01** | 68 (61-76)<br>0.12 |
| Male Gender<br>n (%)<br>*p* value | 515 (64.9%) | 158 (65.3%)<br>0.98 | 207 (63.8%)<br>0.79 | 45 (70.3%)<br>0.32 | 63 (64.9%)<br>1 | 24 (58.5%)<br>0.5 | 102 (63.4%)<br>0.77 | 245 (64.6%)<br>0.21 | 162 (66.9%)<br>0.62 |
| LOS in days<br>Mean (IQR)<br>*p* value | 7.0 (4.0-12.0) | 4.5 (3.1-6.8)**<br>< **0.001** | 7.0 (4.2-11.3)<br>0.87 | 3.3 (2.5-4.9)*<br>< **0.01** | 5.3 (3.3-8.2)*<br>< **0.01** | 6.5 (4.3-9.2)<br>0.67 | 8.3 (4.3-14.7)*<br>**0.03** | 6.0 (4.2-9.2)*<br>**0.01** | 9.8 (6.4-15.9)*<br>< **0.01** |
| + Opioid<br>n (%)<br>*p* value | 413 (52.1%) | 121 (50.2%)<br>0.62 | 220 (67.9%)**<br>< **0.001** | 27 (42.2%)<br>0.16 | 59 (60.8%)<br>0.13 | 24 (58.5%)<br>0.5 | 120 (74.5%)*<br>< **0.01** | 221 (58.3%)*<br>**0.048** | 175 (72.3%)*<br>< **0.01** |
| MME/d Opioid<br>Mean (IQR)<br>*p* value | 4.4 (2-14.8) | 4.8 (1.9-16.4)<br>0.79 | 5.1 (2-14)<br>0.39 | 6.3 (2-14.8)<br>0.52 | 4.6 (3-13.1)<br>0.35 | 4.3 (1.5-11.6)<br>0.68 | 5.9 (1.9-16.9)<br>0.6 | 5.4 (2.2-16.1)<br>0.24 | 5.2 (2-16.3)<br>0.06 |
| + Acetaminophen<br>n (%)<br>*p* value | 102 (12.9%) | 22 (9.1%)<br>0.14 | 39 (12%)<br>0.78 | 4 (6.3%)<br>0.18 | 10 (10.3%)<br>0.58 | 6 (14.6%)<br>0.93 | 20 (12.4%)<br>0.98 | 47 (12.4%)<br>0.9 | 36 (14.9%)<br>0.48 |

In examining opioid use, we found that CLI encounters were more likely to use opioids as compared to the CON group (CLI 67.9% versus CON 52.1%; χ^2^ = 22.8 df = 1; *p* <0.001; chi-squared test) (**Table I**). Additionally, opioid use was more common in gangrene encounters and in both vascular treatment groups (REVASC and AMP) as compared to CON (Gangrene vs. CON: χ^2^ = 26.5 df = 1, *p* < 0.01; REVASC vs. CON: χ^2^ = 15.9 df = 1, *p* = 0.048; AMP vs. CON: χ^2^ = 30.1 df = 1, *p* < 0.01; chi-squared test) (**Table I**). We found no significant differences in MME/d calculated for any of the investigated groups as compared to CON (**Table I**).

Given that acetaminophen is often used as a non-opioid analgesic option for patients undergoing surgical procedures [12], we investigated in-hospital use of acetaminophen between groups. We found that all groups used acetaminophen at similar rates compared to CON with no significant difference in the use of opioids versus acetaminophen across the nine groups (χ^2^ = 5.409, df = 8, *p* = 0.7131; chi-square test) (**Table 1**). These results indicate that individuals in one group were not more likely to be given acetaminophen compared to individuals in another group.

To examine in-patient opioid use patterns in subjects with comorbid DM, PAD and CLI encounters were separated by a diagnosis of DM and demographics for the groups were evaluated. No difference in age or gender was noted between the PAD or CLI groups with or without a diagnosis of DM (PAD comparison χ^2^ = 0.87 df = 1, CLI comparison χ^2^ = 0.44 df = 1; chi-squared test) (**Table 2**). Both PAD +DM and CLI +DM encounters were found to have significantly longer hospital admissions as compared to their respective -DM groups (PAD comparison: *W* = 3853, *p* < 0.01; CLI comparison: *W* = 9548, *p* < 0.01; Wilcoxon test) (**Table 2**). The frequency of narcotic use, MME/d and the frequency of acetaminophen use were not found to be significantly different with a diagnosis of DM (PAD comparison: χ^2^ = 3.46 df =1, CLI comparison: χ^2^ = 0.94 df = 1; chi-squared test) (**Table 2**).

**Table 2.** Institutional data separated by a diagnosis of DM and vascular disease severity (PAD and CLI). Percentage of hospital admissions positive for opioids noted across “+ opioid” row. \**p* < 0.05.

| **Group**<br>n (%) | **PAD-DM**<br>**68 (28.1%)** | **PAD+DM**<br>**174 (71.9%)** | **CLI-DM**<br>**179 (55.2%)** | **CLI+DM**<br>**145 (44.8)** |
| --- | --- | --- | --- | --- |
| **Age in years**<br>Median (IQR)<br>*p* value | 69.5 (63-74) | 69.5 (62-75)<br>0.82 | 68 (62-77) | 67 (60-77)<br>0.40 |
| **Male Gender**<br>n (%)<br>*p* value | 48 (70.6%) | 110 (63.2)<br>0.35 | 111 (62%) | 96 (66.2%)<br>0.51 |
| **LOS in days**<br>Median (IQR)<br>*p* value | 3.4 (2.5-4.9) | 5.3 (3.3-7.5)*<br>**<0.01** | 6 (3.7-9.1) | 8.9 (4.9-15.0)*<br>**<0.01** |
| **+ Opioid**<br>n (%)<br>*p* value | 27 (39.7%) | 94 (54.0%)<br>0.06 | 117 (65.4%) | 103 (71.0%)<br>0.33 |
| **MME/d Opioid**<br>Median (IQR)<br>*p* value | 6.3 (2-14.8) | 4.6 (1.9-15.5)<br>0.65 | 5.9 (2.9-15.8) | 4.6 (1.5-16)<br>0.11 |
| **+ Acetaminophen**<br>n (%)<br>*p* value | 4 (5.9%) | 18 (10.3%)<br>0.4 | 19 (10.6%) | 20 (13.8%)<br>0.48 |

PAD +/-DM and CLI +/-DM were further separated by vascular treatment (REVASC or AMP) and the narcotic use frequency and MME/d were evaluated. Of note, 26 PAD-DM, 56 PAD +DM, 52 CLI -DM, and 36 CLI +DM encounters underwent no intervention (**Table 3**). We found that patients with DM undergoing REVASC did not use narcotics more frequently than their respective -DM groups (PAD REVASC comparison: χ^2^ = 0.0253 df =1; CLI REVASC comparison: χ^2^ = 0.6808 df =1; chi-squared test) (**Table 3**). Additionally, no difference in MME/d was noted for PAD +DM and PAD –DM encounters undergoing REVASC (*W* = 506; Wilcoxon test) and no difference in MME/d was noted for CLI +MD and CLI -DM encounters undergoing REVASC (*W* = 504; Wilcoxon test) (**Table 3**). There was no significant difference in narcotic use frequency between CLI +DM and CLI -DM encounters undergoing AMP (CLI AMP comparison: χ^2^ = 0.24 df =1; chi-squared test) (**Table 3**). MME/d calculated for the CLI +DM encounters undergoing AMP was significantly less than the MME/d calculated for CLI - DM encounters undergoing AMP (CLI REVASC comparison: *W* = 629, *p* = 0.01; Wilcoxon test) (**Table 3**).

**Table 3.** Opioid use for vascular procedure treatment groups (REVASC and AMP) separated by a diagnosis of DM and vascular disease severity. Percentage of hospital admissions positive for opioids noted across “+ opioid” rows. \**p* < 0.05.

| **Group**<br>n (%) | **PAD-DM**<br>**68 (28.1%)** | **PAD+DM**<br>**174 (71.9%)** |  | **CLI-DM**<br>**179 (55.2%)** | **CLI+DM**<br>**145 (44.8)** |
| --- | --- | --- | --- | --- | --- |
| **REVASC** | n=42 | n=103 |  | n=91 | n=23 |
| **+ Opioid**<br>n (%)<br>*p* value | 21 (50%) | 50 (48.5%)<br>1 |  | 59 (64.8%) | 17 (73.9%)<br>.56 |
| **MME/d Opioid**<br>Mean (IQR)<br>*p* value | 3.9 (1.8-12.4) | 5.3 (1.8-17.3)<br>0.82 |  | 5.8 (2.3-19.0) | 8.1 (1.5-14.4)<br>0.98 |
| **AMP** | n=0 | n=15 |  | n=36 | n=86 |
| **+ Opioid**<br>n (%)<br>*p* value | 0 | 10 (66.7%)<br>- |  | 29 (80.6%) | 64 (74.4%)<br>0.62 |
| **MME/d Opioid**<br>Mean (IQR)<br>*p* value | - | 9.6 (3.2-16.1)<br>- |  | 9.1 (4.2-27.6) | 4.2 (1.9-14.1)*<br>**0.01** |

### Outpatient opioid use

During the five-year period, a total of 438 patients (representing 242 PAD and 324 CLI encounters) were admitted to the hospital with a primary diagnosis of either PAD or CLI and retrospectively reviewed for outpatient opioid use. At one month after most recent lower extremity vascular surgery intervention for revascularization or amputation, 238 (54.3%) were no longer taking opioids and 186 (42.5%) continued to take opioids. A total of 14 (3.2%) patients had no follow-up to assess opioid use. The most common opioid prescribed was oxycodone followed by hydrocodone-acetaminophen and hydromorphone. For those using opioids greater than one month, the median use time was three months (IQR 3-8 months). By six months after the most recent lower extremity vascular intervention, 54 (12.3%) patients were still using opioids. By twelve months, 31 (7.1%) patients were still using opioids. By study end, 108 of the 438 (24.6%) patients treated during the five-year study period were deceased. There were no differences in the number of deceased patients taking narcotics and not taking narcotics (*p* =0.38; chi-squared test).

### Post-intervention outpatient opioid use

During a five-year period, 81 of 438 patients (18.5%) were noted to have multiple encounters for lower extremity vascular treatments. These 81 patients underwent a total of 345 life-time vascular procedures with a median of four admissions per patient. Opioid use for these 81 patients was evaluated beyond their most recent hospital admission for vascular intervention. A total of 49 (60.5%) patients were found to be using opioids beyond one month after vascular intervention. 45 of the 49 patients still using opioids had outpatient records available for evaluation beyond one month. These 45 patients used opioids for a median of 4 months (IQR 2-8 months). Nine (11%) patients were using opioids beyond one year of their last vascular surgery. A total of 21 (25.9%) patients died during the study period with no overt deaths attributable to opioid overdose. There was no statistical significance in the number of deaths for patients using opioids beyond one month after their last vascular surgery as compared to the number of deaths for patients not using opioids (12 (14.8%) versus 8 (9.9%), *p* = 0.98; chi squared test). Of note, 10 of the 81(12.3%) patients were transitioned to tramadol and taking it beyond one month of most recent vascular intervention. Three patients on tramadol (33%) were deceased at the end of the study.

We next examined the association between vascular procedures and opioid use beyond one month of procedure. A chi-square test was conducted to analyze the relationship between the type of vascular procedure performed and opioid use rates. The analysis included data from 45 patients who underwent 10 different vascular procedures and who had outpatient records available. Our results show a statistically significant association between vascular procedure type and opioid use rates (χ^2^ = 59.89, df = 9, *p* < 0.0001; chi-square test) (**Figure 1**). These findings indicate that the likelihood of opioid use varied significantly depending upon the type of vascular procedure performed. However, it’s important to note that these results may also be influenced by the number of procedures undergone per patient.

**Figure 1.**
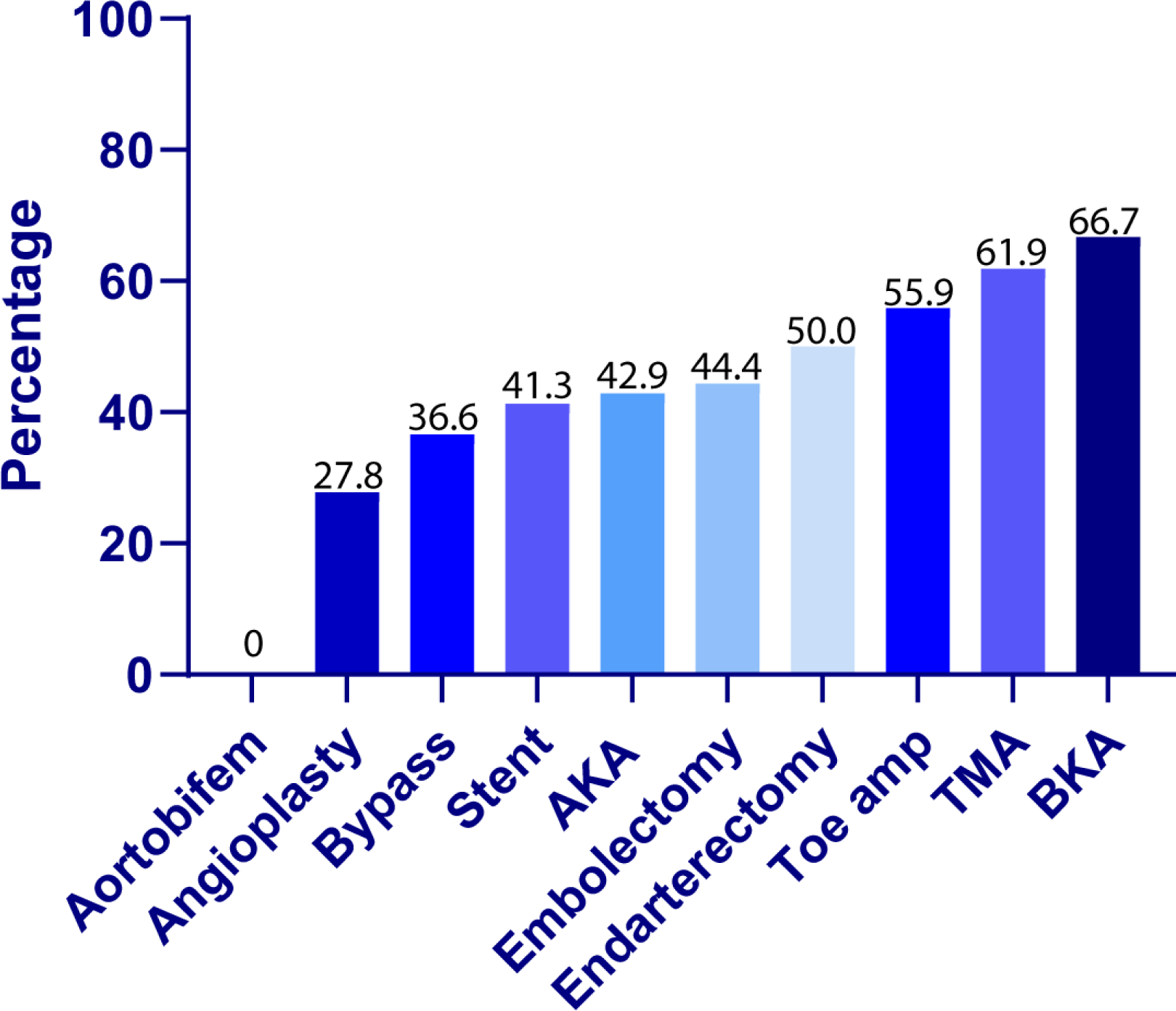
The percentage of patients having undergone each procedure type still using opioids beyond 1 month of last vascular surgery. Corresponding *P* values depicted at base of the bars. Abbreviations: Aortobifem = aortobifemoral bypass surgery; Bypass = peripheral artery bypass surgery; Stent = peripheral artery stent placement; AKA = above knee amputation; TMA = transmetatarsal amputation; BKA = below knee amputation.

Finally, we compared patients taking opioid beyond one month of last vascular intervention to those not taking opioid for total number of vascular procedures performed. Patients continuing to take opioids were not more likely to have more procedures performed as compared to those patients no longer taking opioids (3.9 procedures versus 4.1 procedures, *p* = 0.74; chi-squared test). For the patients continuing to take opioid beyond one month of last vascular intervention, no correlation between the number of vascular surgeries performed on the patient was observed with the number of months the patient was reported to have been taking opioid (*r* = -0.0052, *p* = 0.97; Pearson’s correlation coefficient).

## DISCUSSION

This institutional retrospective review of opioid use in patients affected with lower extremity arterial disease confirms that for in-hospital encounters with increasing disease severity (CLI versus PAD, CLI versus CON, and gangrene versus CON group), the frequency of opioid use was increased. When evaluating lower extremity disease severity as associated with specific signs/symptoms (i.e., claudication, rest pain, ulceration and gangrene), this study did not find sequential increases in the frequency of opioid use with progressive disease until gangrene was present. Notably, the amount of opioid used to treat patients with CLI and gangrene was not significantly more. There was an increased frequency of opioid use for hospital encounters to treat lower extremity arterial disease (REVASC and AMP) as compared to the CON group. However, the amount of opioid used was not significantly greater. These findings suggest that patients with CLI, with gangrene and or undergoing a vascular procedure (REVASC and AMP) are not exposed to increased amounts of opioid during their hospitalization. A diagnosis of DM did not seem to affect the frequency of narcotic usage, however, encounters for CLI +DM patients undergoing AMP received significantly smaller doses of opioid at the same frequency. This finding can likely be attributed to a sicker, more fragile patient population for which limited opioid dose is preferred to maintain hemodynamic stability during the perioperative period. Moreover, the rate of acetaminophen use for all these populations is uniformly low as compared to the rate of opioid use.

Outpatient opioid usage for all patients evaluated, undergoing single and multiple interventions, at one month and one year was 42.5% and 7.1%, respectively. Approximately 60% of patients undergoing multiple vascular interventions were found to be using opioids beyond one month of their most recent vascular surgery intervention which was significantly increased as compared to those undergoing single vascular procedures. Only 11% of these patients having undergone multiple vascular procedures were still using opioids at one year and there was no statistical significance as compared to the number of patients treated with a single vascular intervention taking opioids one year out. These findings suggest that the frequency of opioid usage in the outpatient post-operative period after multiple vascular interventions might be increased initially, but the difference was not maintained beyond one year.

While one quarter of the patients undergoing multiple vascular procedures are deceased at the study end, none of these patients were deceased from opioid overdose. Patients undergoing multiple procedures were more likely to be using opioids for more than one month after recent vascular surgery intervention. Patients having undergone multiple vascular procedures and using opioids beyond one month of their most recent vascular surgery intervention were not found to have an increased number of vascular surgery procedures performed as compared to other patients having undergone multiple vascular procedures, not using opioids. This suggests that increased procedure number does not necessarily render the patient more susceptible to opioid dependence. No correlation between vascular procedure number and number of months of opioid use could be shown.

Similar to Itoga, NK et al, our findings suggest that increased lower extremity arterial disease severity with progression to CLI was associated with more frequent opioid use during the hospital encounter, however, such patients were not treated with higher amounts of opioid [13]. And as Itoga NK et al suggests, the frequency of opioid use during hospital encounters increases with REVASC. AMP also showed an increased rate of opioid usage during hospital encounters, even as compared to REVASC. Because of data limitations, we were not able to examine opioid use frequency or dose amounts in patients admitted for multiple hospital encounters.

Additionally, we found that the rate of persistent opioid use (beyond one month) in patients undergoing multiple vascular procedures was comparable to previous reports that investigated chronic opioid use history in patients undergoing vascular interventions within the one to three months post-operative period [14,15]. Notably, half of the patients treated with an entirely endovascular approach were persistent opioid users showing that endovascular only revascularizations are not the answer for reducing chronic narcotic use in this patient population [16]. This study also shows that many of those patients can be weaned successfully off opioids by one year after the most recent vascular procedure with long term opioid prevalence similar to other surgical specialties [10]. Even though patients undergoing multiple vascular surgery interventions do not appear to have a predilection for developing opioid addiction, vascular surgery prescribers should be aware that there are some procedures that are more likely to result in long-term opioid use. Interestingly, others have suggested that prescribers are often sending patients home with prescriptions that are for higher amounts of narcotic per day and do not reflect the actual opioid usage during hospitalization [17]. Based on encounter dosing, our study suggests that patients should not be receiving more than 5 mg of oxycodone with 2-3 tablets per day at the time of discharge. We could find no studies suggesting benefit or lack of benefit in treating PAD at any severity with acetaminophen.

Study limitations include single hospital/institution data with limited numbers representing a limited patient population and limited surgical team. Our analysis also revealed that ICD codes applied to represent disease severity might not be the most accurate as patients with peripheral arterial disease, without critical limb ischemia, were given CPT codes designating they underwent amputation. With numerous ICD codes to describe to the patient presentation, vascular surgeons should attempt to be as specific as possible with diagnosis.

Finally, a better understanding of the pain associated with vascular ischemia is desperately needed. While revascularization, if possible, is the most important tenet of vascular surgery treatment, patients will suffer pain until an accurate diagnosis is made, appropriate treatment can be performed as well as initially in the post-operative period. Focus within the vascular surgery community should be directed at developing pain medications specifically foiling the ischemic response.

## CONCLUSIONS

In conclusion, we find that patients with CLI and gangrene as well as those undergoing vascular treatment have a greater frequency of opioid use during hospital encounters as compared those patients with less severe disease and undergoing conservative management, respectively. However, these findings do not equate to higher doses of opioid used during hospitalization. These data suggest that healthcare providers are opting for smaller, more frequent doses of opioid to maintain a consistent level of pain relief, while minimizing the risk of adverse side effects, including respiratory depression. Finally, we find that patients undergoing multiple vascular procedures are not more likely to be using opioid long-term (at one year) as compared to those patients treated with single vascular procedures.

## Data Availability

All data produced in the present work are contained in the manuscript

---

**Supplemental Table 1.** ICD codes designating patients with lower extremity atherosclerotic disease or lower extremity diabetic peripheral angiopathy included (columns listed PAD codes and CLI codes) and ICD codes excluded for upper extremity atherosclerotic disease or unspecified.

| **PAD codes** | **CLI codes** | **Codes Excluded for Unspecified Limb or Arm** |
| --- | --- | --- |
| I70.2 | I70.22 | I70.208 |
| I70.20 | I70.221 | I70.209 |
| I70.201 | I70.222 | I70.218 |
| I70.202 | I70.223 | I70.219 |
| I70.203 | I70.23 | I70.228 |
| I70.21 | I70.231 | I70.229 |
| I70.211 | I70.232 | I70.25 |
| I70.212 | I70.233 | I70.268 |
| I70.213 | I70.234 | I70.269 |
| I70.29 | I70.235 | I70.298 |
| I70.291 | I70.238 | I70.299 |
| I70.292 | I70.239 | I70.308 |
| I70.293 | I70.24 | I70.309 |
| I70.3 | I70.241 | I70.318 |
| I70.30 | I70.242 | I70.319 |
| I70.301 | I70.243 | I70.328 |
| I70.302 | I70.244 | I70.329 |
| I70.303 | I70.245 | I70.35 |
| I70.31 | I70.248 | I70.368 |
| I70.311 | I70.249 | I70.369 |
| I70.312 | I70.26 | I70.398 |
| I70.313 | I70.261 | I70.399 |
| I70.39 | I70.262 | I70.408 |
| I70.391 | I70.263 | I70.409 |
| I70.392 | I70.32 | I70.418 |
| I70.393 | I70.321 | I70.419 |
| I70.40 | I70.322 | I70.428 |
| I70.401 | I70.323 | I70.429 |
| I70.402 | I70.33 | I70.45 |
| I70.403 | I70.331 | I70.468 |
| I70.41 | I70.332 | I70.469 |
| I70.411 | I70.333 | I70.498 |
| I70.412 | I70.334 | I70.499 |
| I70.413 | I70.335 | I70.508 |
| I70.49 | I70.338 | I70.509 |
| I70.491 | I70.339 | I70.518 |
| I70.492 | I70.34 | I70.519 |
| I70.493 | I70.341 | I70.528 |
| I70.50 | I70.342 | I70.529 |
| I70.501 | I70.343 | I70.55 |
| I70.502 | I70.344 | I70.568 |
| I70.503 | I70.345 | I70.569 |
| I70.51 | I70.348 | I70.598 |
| I70.511 | I70.349 | I70.599 |
| I70.512 | I70.36 | I70.608 |
| I70.513 | I70.361 | I70.609 |
| I70.59 | I70.362 | I70.618 |
| I70.591 | I70.363 | I70.619 |
| I70.592 | I70.42 | I70.628 |
| I70.593 | E11. 52 | I70.629 |
| I70.6 | I70.421 | I70.65 |
| I70.60 | I70.422 | I70.668 |
| I70.601 | I70.423 | I70.669 |
| I70.602 | I70.43 | I70.698 |
| I70.603 | I70.431 | I70.699 |
| I70.61 | I70.432 | I70.708 |
| I70.611 | I70.433 | I70.709 |
| i70.612 | I70.434 | I70.718 |
| I70.613 | I70.435 | I70.719 |
| I70.69 | I70.438 | I70.728 |
| I70.691 | I70.439 | I70.729 |
| I70.692 | I70.44 | I70.75 |
| I70.693 | I70.441 | I70.768 |
| I70.7 | I70.442 | I70.769 |
| I70.70 | I70.443 | I70.798 |
| I70.701 | I70.444 | I70.799 |
| I70.702 | I70.445 |  |
| I70.703 | I70.448 |  |
| I70.71 | I70.449 |  |
| I70.711 | I70.46 |  |
| I70.712 | I70.461 |  |
| I70.713 | I70.462 |  |
| I70.79 | I70.463 |  |
| I70.791 | I70.52 |  |
| I70.792 | I70.521 |  |
| I70.793 | I70.522 |  |
| I70.8 | I70.523 |  |
| I70.9 | I70.53 |  |
| I70.90 | I70.531 |  |
| I70.91 | I70.532 |  |
| I70.92 | I70.533 |  |
| I73.9 | I70.534 |  |
| I73.8 | I70.535 |  |
| I73.89 | I70.538 |  |
| Z86.79 | I70.539 |  |
| E08. 51 | I70.54 |  |
| E08. 59 | I70.541 |  |
| E09. 51 | I70.542 |  |
| E09. 59 | I70.543 |  |
| E10. 51 | I70.544 |  |
| E10. 59 | I70.545 |  |
| E11. 51 | I70.548 |  |
| E11. 59 | I70.549 |  |
| E13. 51 | I70.56 |  |
| E13. 59 | I70.561 |  |
|  | I70.562 |  |
|  | I70.563 |  |
|  | I70.62 |  |
|  | I70.621 |  |
|  | E13. 52 |  |
|  | I70.622 |  |
|  | I70.623 |  |
|  | I70.63 |  |
|  | I70.631 |  |
|  | I70.632 |  |
|  | I70.633 |  |
|  | I70.634 |  |
|  | I70.635 |  |
|  | I70.638 |  |
|  | I70.639 |  |
|  | I70.64 |  |
|  | I70.641 |  |
|  | I70.642 |  |
|  | I70.643 |  |
|  | I70.644 |  |
|  | I70.645 |  |
|  | I70.648 |  |
|  | I70.649 |  |
|  | I70.66 |  |
|  | I70.661 |  |
|  | I70.662 |  |
|  | I70.663 |  |
|  | I70.72 |  |
|  | I70.721 |  |
|  | I70.722 |  |
|  | I70.723 |  |
|  | I70.73 |  |
|  | I70.731 |  |
|  | I70.732 |  |
|  | I70.733 |  |
|  | I70.734 |  |
|  | I70.735 |  |
|  | I70.738 |  |
|  | I70.739 |  |
|  | I70.74 |  |
|  | I70.741 |  |
|  | I70.742 |  |
|  | I70.743 |  |
|  | I70.744 |  |
|  | I70.745 |  |
|  | I70.748 |  |
|  | I70.749 |  |
|  | I70.76 |  |
|  | I70.761 |  |
|  | I70.762 |  |
|  | I70.763 |  |
|  | E08. 52 |  |
|  | E09. 52 |  |
|  | E10. 52 |  |

**Supplemental Table 2.**
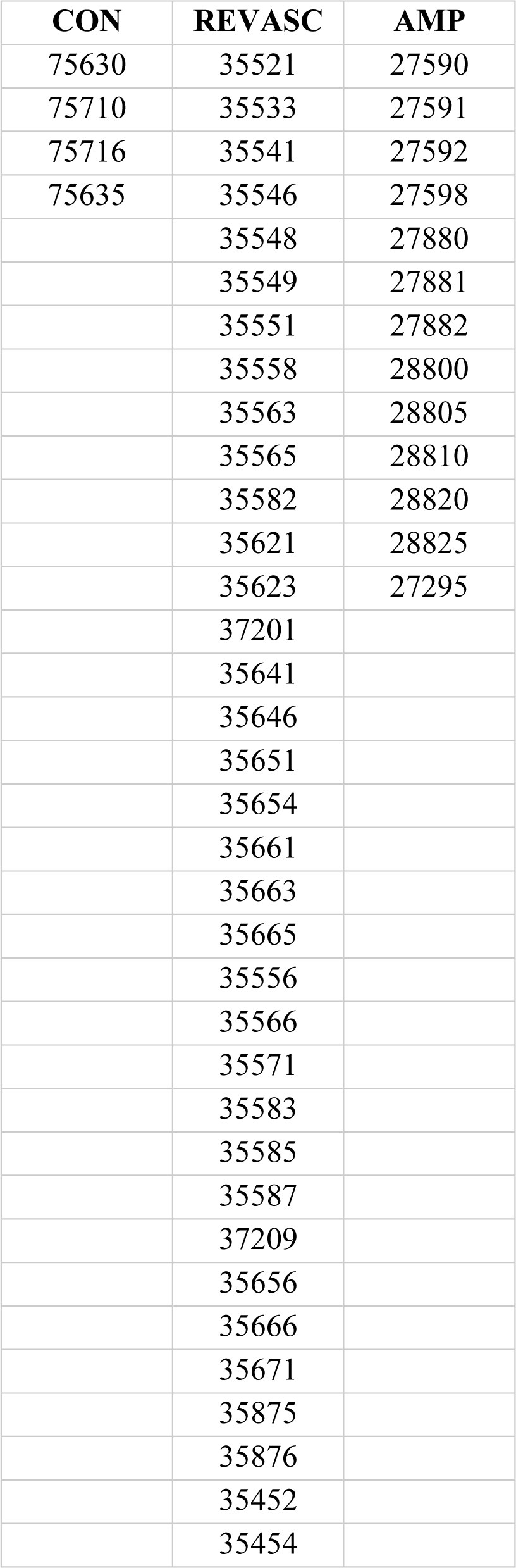
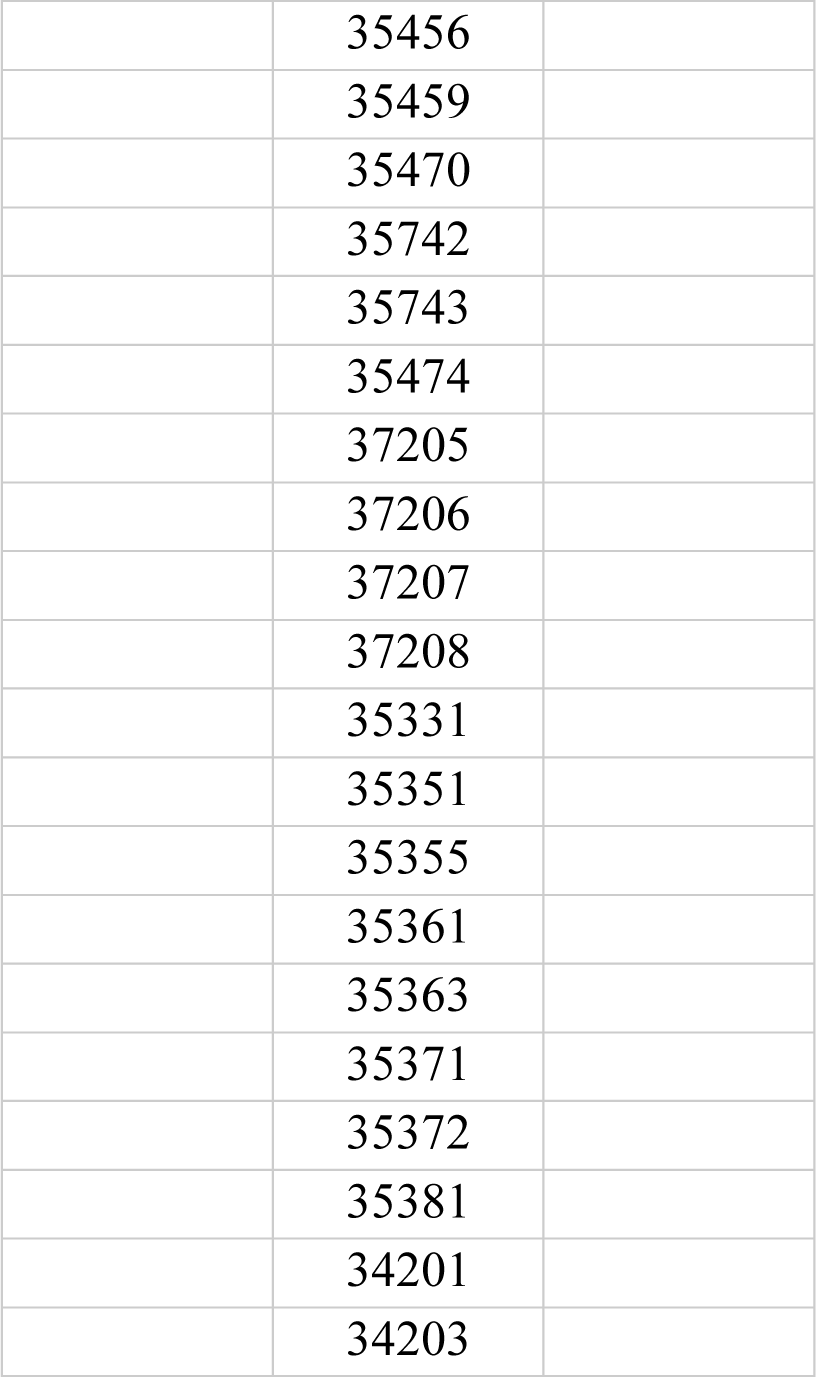
CPT codes designating diagnostic imaging procedures for CON, procedures performed for REVASC, and procedures performed for AMP.

| CON | REVASC | AMP |
| --- | --- | --- |
| 75630 | 35521 | 27590 |
| 75710 | 35533 | 27591 |
| 75716 | 35541 | 27592 |
| 75635 | 35546 | 27598 |
|  | 35548 | 27880 |
|  | 35549 | 27881 |
|  | 35551 | 27882 |
|  | 35558 | 28800 |
|  | 35563 | 28805 |
|  | 35565 | 28810 |
|  | 35582 | 28820 |
|  | 35621 | 28825 |
|  | 35623 | 27295 |
|  | 37201 |  |
|  | 35641 |  |
|  | 35646 |  |
|  | 35651 |  |
|  | 35654 |  |
|  | 35661 |  |
|  | 35663 |  |
|  | 35665 |  |
|  | 35556 |  |
|  | 35566 |  |
|  | 35571 |  |
|  | 35583 |  |
|  | 35585 |  |
|  | 35587 |  |
|  | 37209 |  |
|  | 35656 |  |
|  | 35666 |  |
|  | 35671 |  |
|  | 35875 |  |
|  | 35876 |  |
|  | 35452 |  |
|  | 35454 |  |
|  | 35456 |  |
|  | 35459 |  |
|  | 35470 |  |
|  | 35742 |  |
|  | 35743 |  |
|  | 35474 |  |
|  | 37205 |  |
|  | 37206 |  |
|  | 37207 |  |
|  | 37208 |  |
|  | 35331 |  |
|  | 35351 |  |
|  | 35355 |  |
|  | 35361 |  |
|  | 35363 |  |
|  | 35371 |  |
|  | 35372 |  |
|  | 35381 |  |
|  | 34201 |  |
|  | 34203 |  |

## REFERENCES CITED

1. Fowkes FG, Aboyans V, Fowkes FJ, McDermott MM, Sampson UK, Criqui MH: Peripheral artery disease: epidemiology and global perspectives. Nat Rev Cardiol. 2017, 14:156–170. 10.1038/nrcardio.2016.179

2. Clair D, Shah S, Weber J: Current state of diagnosis and management of critical limb ischemia. Curr Cardiol Rep. 2012, 14:160–170. 10.1007/s11886-012-0251-4

3. Ikhlas M, Atherton N: 2023 Vascular Reperfusion Injury. StatPearls.

4. Richardson C, Glenn S, Horgan M, Nurmikko T: A Prospective Study of Factors Associated With the Presence of Phantom Limb Pain Six Months After Major Lower Limb Amputation in Patients With Peripheral Vascular Disease. The Journal of Pain. 2007, 8:793–801. 10.1016/j.jpain.2007.05.007

5. Richardson C, Glenn S, Nurmikko T, Horgan M: Incidence of Phantom Phenomena Including Phantom Limb Pain 6 Months After Major Lower Limb Amputation in Patients With Peripheral Vascular Disease. The Clinical journal of pain. 2006, 22:353–358. 10.1097/01.ajp.0000177793.01415.bd

6. Drożdżal S, Lechowicz K, Szostak B, et al.: Kidney damage from nonsteroidal anti-inflammatory drugs-Myth or truth? Review of selected literature. Pharmacol Res Perspect. 2021, 9:e00817. 10.1002/prp2.817

7. Fraser K, Raju I: Anaesthesia for lower limb revascularization surgery. BJA Education. 2014, 15:225–230. 10.1093/bjaceaccp/mku042

8. Ballantyne JC, Shin NS: Efficacy of Opioids for Chronic Pain: A Review of the Evidence. The Clinical journal of pain. 2008, 24:469–478. 10.1097/AJP.0b013e31816b2f26

9. Manhapra A, MacLean RR, Rosenheck R, Becker WC: Are opioids effective analgesics and is physiological opioid dependence benign? Revising current assumptions to effectively manage long-term opioid therapy and its deprescribing. British Journal of Clinical Pharmacology. n/a. 10.1111/bcp.15972

10. Jiang X, Orton M, Feng R, et al.: Chronic Opioid Usage in Surgical Patients in a Large Academic Center. Ann Surg. 2017, 265:722–727. 10.1097/sla.0000000000001780

11. Dubois L, McClure JA, Vogt K, Welk B, Clarke C: Association Between Complications after Vascular Surgery and Prolonged Postoperative Opioid Use. Ann Vasc Surg. 2024, 98:274–281. 10.1016/j.avsg.2023.08.008

12. Anderson M, Hallway A, Brummett C, Waljee J, Englesbe M, Howard R: Patient-Reported Outcomes After Opioid-Sparing Surgery Compared With Standard of Care. JAMA Surg. 2021, 156:286–287. 10.1001/jamasurg.2020.5646

13. Itoga NK, Sceats LA, Stern JR, Mell MW: Association of opioid use and peripheral artery disease. J Vasc Surg. 2019, 70:1271–1279.e1271. 10.1016/j.jvs.2018.12.036

14. Howard R, Albright J, Englesbe M, Osborne N, Henke P: Opioid use in patients with peripheral arterial disease undergoing lower extremity bypass. J Vasc Surg. 2022, 75:998–1007. 10.1016/j.jvs.2021.08.104

15. Velazquez-Ramirez G, Krebs J, Stafford JM, et al.: Prevalence of chronic opioid use in patients with peripheral arterial disease undergoing revascularization. J Vasc Surg. 2022, 75:186–194. 10.1016/j.jvs.2021.07.236

16. Brown CS, Osborne NH, Hu HM, et al.: Endovascular surgery is not protective against new persistent opioid use development compared to open vascular surgery. Vascular. 2022, 30:728–738. 10.1177/17085381211024514

17. Camazine MN, Rountree KM, Smith JB, Bath J, Vogel TR: Opioid utilization after lower extremity amputation for peripheral vascular disease and discharge prescribing recommendations. Vascular. 2023, 31:954–960. 10.1177/17085381221097163

